# S gene dropout patterns in SARS-CoV-2 tests suggest spread of the H69del/V70del mutation in the US

**DOI:** 10.1101/2020.12.24.20248814

**Authors:** Nicole L. Washington, Simon White, Kelly M. Schiabor Barrett, Elizabeth T. Cirulli, Alexandre Bolze, James T. Lu

**Author notes:** Corresponding Author: Nicole Washington.

## Abstract

Recently, multiple novel strains of SARS-CoV-2 have been found to share the same deletion of amino acids H69 and V70 in the virus S gene. This includes strain B.1.1.7 / SARS-CoV-2 VUI 202012/01, which has been found to be more infectious than other strains of SARS-CoV-2, and its increasing presence has resulted in new lockdowns in and travel restrictions leaving the UK. Here, we analyze 2 million RT-PCR SARS-CoV-2 tests performed at Helix to identify the rate of S gene dropout, which has been recently shown to occur in tests from individuals infected with strains of SARS-CoV-2 that carry the H69del/V70del mutation. We observe a rise in S gene dropout in the US starting in early October, with 0.25% of our daily SARS-CoV-2-positive tests exhibiting this pattern during the first week. The rate of positive samples with S gene dropout has grown slowly over time, with last week exhibiting the highest level yet, at 0.5%. Focusing on the 14 states for which we have sufficient sample size to assess the frequency of this rare event (n>1000 SARS-CoV-2-positive samples), we see a recent expansion in the Eastern part of the US, concentrated in MA, OH, and FL. However, we cannot say from these data whether the S gene dropout samples we observe here represent the B.1.1.7. strain. Only with an expansion of genomic surveillance sequencing in the US will we know for certain the prevalence of the B.1.1.7 strain in the US.

---

Since the onset of the COVID-19 pandemic, there has been concern about novel SARS-CoV-2 mutations emerging that are more transmissible. Recently, multiple novel strains of SARS-CoV-2 have been found to share the same deletion of amino acids H69 and V70 in the virus S gene. At least some of these strains are hypothesized to have increased transmissibility, and some have been found to infect both minks and humans^1,2^. This includes strain B.1.1.7 / SARS-CoV-2 VUI 202012/01, which has been found to be more infectious than other strains of SARS-CoV-2, and its increasing presence has resulted in new lockdowns in and travel restrictions leaving the UK ^3–5^. The B.1.1.7 strain has not yet been found in the United States, but viral sequencing of SARS-CoV-2 for surveillance purposes has been very limited and performed on only a relatively small number of samples.

The Helix^®^ COVID-19 Test includes amplification of three viral genes: Orf1ab, N and the S genes^6^. A recent preprint has shown that deletion of H69 and V70 can be characterized by a dropout of the S gene in standard and widely used RT-PCR SARS-CoV-2 tests^7^. This analysis technique can therefore serve as a proxy to detect infections driven by these newly emerging sequence variants. Given the new information about the link between S gene dropout and the H69del/V70del mutation, together with the concern over increased transmissibility of the B.1.1.7 strain, we examined the prevalence of S gene dropout in 2 million samples from across the US tested at Helix.

We observe a rise in S gene dropout starting in early October, with 0.25% of our daily SARS-CoV-2-positive tests exhibiting this pattern during the first week (Figure 1). The rate of positive samples with S gene dropout has grown slowly over time, with last week exhibiting the highest level yet at 0.5% of SARS-CoV-2-positive tests that are consistent with the H69del/V70del variant.

**Figure 1.**
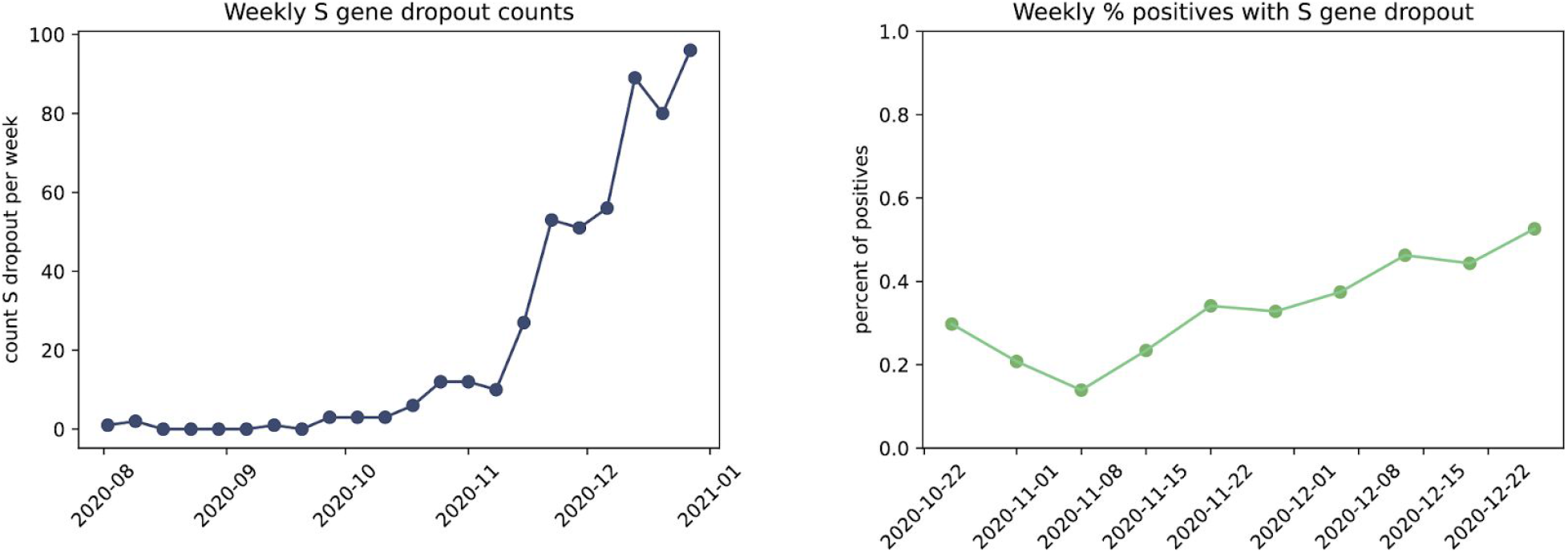
Weekly observations of S gene dropout. The raw number of tests per week with S gene dropout (left), as well as normalized by the number of positive tests (right), indicates that the presence of the H69del/V70del or potentially other S gene variants is increasing in our tested population.

Since we test samples from all 50 states in our laboratory, we examined the nationwide distribution of these S gene dropout positive samples as well as state-level trends (Figure 2). We have observed S gene dropout positive samples in 19 states so far. Focusing on the 14 states for which we have sufficient sample size to assess the frequency of this rare event (n>1000 SARS-CoV-2-positive samples), we see a recent expansion in the Eastern part of the US, concentrated in MA, OH, and FL.

**Figure 2.**
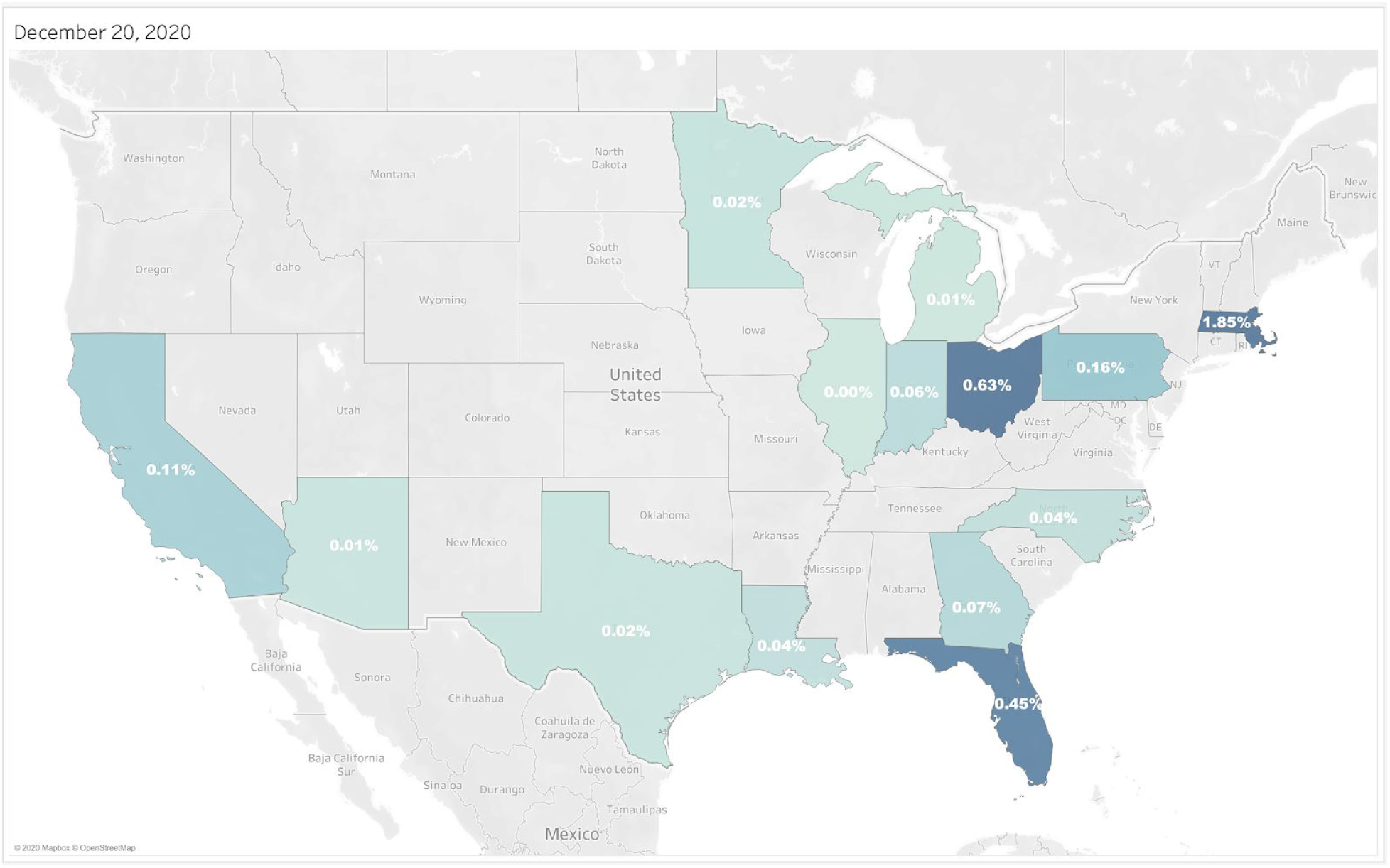
US cumulative distribution of S gene dropout-positive SARS-CoV-2 tests as of Dec 20, 2020. Values indicate percent of positive tests with S gene dropout. To ensure that low sample sizes do not skew frequencies, only states with more than 1,000 SARS-CoV-2 positive tests are shown. States with insufficient data are shown in gray. Additional states with S gene dropout, but with <1K positives in our dataset thus far are: AL, ME, NH, NY, RI, VA. An animated graphic showing the percent of positive samples over time can be found at https://blog.helix.com/sars-cov2_uk-variant/.

As mentioned above, the H69del/V70del variant has been reported in many viral strains available from Nextstrain, which depicts SARS-CoV-2 sequence evolution for epidemiological surveillance^8^. Yet, as of today, only 25 individuals with this variant have been sequenced in the US since March, with all but three found prior to July (when we began testing at Helix)^9^. The rise in S gene dropout in our positive samples in early October is consistent with the rise of H69del/V70del variation globally in recent weeks, including the B.1.1.7 strain that is taking hold in the UK.

B.1.1.7harbors several variants, including the H69del/V70del, but it is unknown which are causing the rise in transmission. We cannot say if the S gene dropout samples we observe here represent the B.1.1.7. strain which harbors the N501Y variant that is said to bind the virus to human cells more tightly, nor P681H that likely affects biological function. Given the exceptionally low fraction of positives that we observe with this variant today, only with an expansion of genomic surveillance sequencing in the US will we know for certain if the B.1.1.7 strain has arrived in the US.

## Limitations

- Following the conservative approach from Bal et al.^7^, for this analysis we only analyzed positive samples with strong amplification of the N gene (Ct < 25), and dropout samples with absolutely no S gene detected. Because some samples may have lower viral titer, and therefore result in a weaker amplification signal, we are not analyzing all positive samples here. Therefore, the final fraction of dropouts may differ slightly.
- Since samples are deidentified prior to analysis, and some individuals may test more than once, there may be some duplicate individuals that could cause deviation from the true population fraction
- Testing per state does not reflect the population distribution of the US, and therefore some states without S gene dropout may be false negatives.
- As we do not store samples longitudinally, we are unable to perform viral sequencing to confirm that our S gene dropouts are indeed deletions of amino acids H69 and V70.
- However, the method used here showed 100% concordance between observed S gene dropout and deletion in viral sequence in the original manuscript.
- Without sequencing, we cannot distinguish between the different viral lineages that carry the H69del/V70del variant. This variant is present in at least three major clades since Oct 2020, including B.1.1.7.

## Data Availability

Please contact authors for information about data availability.

## Notes

### Competing Interest Statement

All authors are employees of Helix.

### Funding Statement

None

### Author Declarations

WCG Institutional Review Board (WCGIRB) approved a request for a waiver for this study under protocol 20203438.

